# Effectiveness of Autumn 2023 COVID-19 vaccination and residual protection of prior doses against hospitalisation in England, estimated using a test-negative case-control study

**DOI:** 10.1101/2024.03.28.24305030

**Authors:** Freja Cordelia Møller Kirsebom, Julia Stowe, Jamie Lopez Bernal, Alex Allen, Nick Andrews

**Affiliations:** UK Health Security Agency, London, United Kingdom; NIHR Health Protection Research Unit in Vaccines and Immunisation, London School of Hygiene and Tropical Medicine, London, United Kingdom; NIHR Health Protection Research Unit in Respiratory Infections, Imperial College London, United Kingdom

**Keywords:** COVID-19, vaccine effectiveness, immunisation, observational study, test-negative case-control

## Abstract

**Introduction:** The last COVID-19 vaccine offered to all adults in England became available from November 2021. The most recent booster programme commenced in September 2023. Bivalent BA.4-5 or monovalent XBB.1.5 boosters were given. During the study period, the JN.1 variant became dominant in England.

**Methods:** Vaccine effectiveness against hospitalisation was estimated throughout using the test-negative case-control study design where positive PCR tests from hospitalised individuals are cases and comparable negative PCR tests are controls. Multivariable logistic regression was used to assess vaccine effectiveness against hospitalisation with the test result as the outcome, vaccination status as the primary exposure variable of interest and confounder adjustment.

**Results:** There was no evidence of residual protection for boosters given as part of previous campaigns. There were 28,916 eligible tests included to estimate the effectiveness of the autumn 2023 boosters in those aged 65 years and older. VE peaked at 50.6% (95% CI: 44.2-56.3%) after 2-4 weeks, followed by waning to 13.6% (95% CI: -11.7-33.2%). Estimates were generally higher for the XBB.1.5 booster than the BA.4-5 booster, but this difference was not statistically significant. Point estimates were highest against XBB sub-lineages. Effectiveness was lower against both JN.1 and EG.5.1 variants with confidence intervals non-overlapping with the effectiveness of the XBB sub-lineages at 2-4 weeks for EG.5.1 where VE was 44.5% (95% CI: 20.2-61.4%) and at 5-9 weeks for JN.1 where VE was 26.4% (95%CI: -3.4-47.6%).

**Conclusions:** The recent monovalent XBB.1.5 and bivalent BA.4-5 boosters provided comparable and good protection against hospitalisation, however there was evidence of lower VE against hospitalisation of these boosters against JN.1.

## Introduction

COVID-19 continues to cause waves of disease, largely driven by the emergence of novel variants and sub-lineages. Although vaccines continue to be updated to better target current variants, the effectiveness of booster vaccines against hospitalisation wanes (1-4). As such, many countries, including England, have opted for regular booster programmes to protect the most vulnerable from severe disease. In England, the XBB-related Omicron sub-lineages were dominant throughout most of 2023 (5). Sub-lineages of BA.2.86 first emerged in during summer 2023 with mutations facilitating high ACE2 binding affinity, although no substantial humoral immune escape or growth advantage was detected (6). JN.1 (BA.2.86.1.1) has an additional receptor binding domain mutation (L455S) and neutralisation studies have found JN.1 has significant enhanced immune escape as compared with BA.2.86 and XBB.1.5 (6). JN.1 was first detected in the United States (US) in September and by mid-December many countries, including England, were detecting an increase in sequenced cases (7). By late January, JN.1 was the dominant sub-lineage in England, accounting for 85% of sequenced cases (5).

As part of the COVID-19 vaccination programme in England, all adults in England were initially offered a primary course (consisting of two vaccine doses, or three doses in the immunosuppressed) and a booster dose, all based on the ancestral strain. The last vaccine offered to all adults in England became available from November 2021 (8). Since, there have been booster vaccination programmes each spring and autumn, targeting risk groups based primarily on age and clinical risk status (9-11). The vaccine type offered has varied as manufacturers have updated formulations to target emergent variants. Adults aged 50 to 64 years of age in England were last offered a booster during the autumn 2022 programme, in which bivalent BA.1 mRNA vaccines were offered to all adults aged 50 years or over and clinically vulnerable people. As of March 2024, the most recent booster programme in England commenced in early September 2023 (12). Adults aged 65 years and older, residents of care homes for older adults, carers and those in a clinical group were offered either a bivalent BA.4-5 booster (based on the ancestral strain and the BA.4-5 sub-lineages) or a monovalent XBB.1.5 booster (12). The roll-out of the BA.4-5 boosters started a few weeks prior to the roll-out of the XBB.1.5 vaccine. Those aged 75 years and older were also eligible for boosters in spring 2022 and most recently in spring 2023 where the Sanofi/GSK AS03-adjuvanted monovalent beta variant vaccine and mRNA bivalent BA.4-5 vaccines were recommended (10).

The aim of this study was to use national level electronic healthcare datasets to estimate the vaccine effectiveness (VE) of the autumn 2023 bivalent BA.4-5 and monovalent XBB.1.5 boosters against hospitalisation. In addition, the residual protection of prior booster doses in the study period was assessed, as well as a variant specific analysis for the autumn 2023 boosters.

## Materials and Methods

### Study design

Vaccine effectiveness against hospitalisation was estimated throughout using the test-negative case-control (TNCC) study design, as previously described (1, 2, 13, 14) where positive PCR tests from hospitalised individuals are cases and negative PCR tests from hospitalised individuals are the controls. The study period was from 4^th^ September 2023 to 21^st^ January 2024, with data extracted on 12^th^ February 2024.

### Data Sources

#### COVID-19 Testing Data

Positive and negative PCR tests undertaken in hospital settings were identified from England’s national COVID-19 testing data. Negative tests within 7 days of a previous negative test and within 21 days of a subsequent positive test were excluded. All tests within 90 days of a previous positive test were excluded as these likely represent the same episode. The date of an individual’s most recent prior positive test was also identified (PCR and self-reported lateral flow tests). Individuals contributed a maximum of one negative test, selected at random. Further details on the reasons for these exclusions are described previously (1, 14). In England, a subset of positive PCR tests undergo whole genome sequencing. Tests with sequencing information associated were included in the variant analysis. Where subsequent tests within 14 days included sequencing information, this was used to enrich variant status.

### Immunisation Information System (IIS)

The testing data were linked to the UKHSA IIS; a national vaccine register (formally the national immunisation management system (NIMS)) containing demographic information on the whole population of England registered with a GP, used to record all COVID-19 vaccination (15), using combinations of the unique individual NHS number, date of birth, surname, first name, and postcode using deterministic linkage. IIS was accessed for dates of vaccination and manufacturer, sex, date of birth, ethnicity, care home residency and residential address. Care home status is provided by the NHS and included those being resident in a care home as of March 2023. Postcodes were used to determine index of multiple deprivation (IMD) quintile (small area measures of relative deprivation based on postcode) and NHS region. Data on clinical risk group status (those identified as requiring an autumn 2023 booster by NHS CaaS (Cohorting as a Service) (16)) were also extracted. Booster doses were classified based on SNOMED coding and timing of vaccination. The following individuals were excluded from the main analyses; those who had received only one dose, those with less than 12 weeks between the latest dose and the dose prior, those who received a vaccine by a manufacturer other than those offered. Additionally, those who were unvaccinated were excluded from all analyses of incremental VE where the comparator group was those who had received at least two prior doses.

### Hospital Admission Data

Secondary Uses Service (SUS) is the national electronic database of hospital admissions that provides timely updates of ICD-10 codes for completed hospital stays for all NHS hospitals in England (17). SUS is an administrative database used by commissioners and providers of NHS-funded care for purposes including healthcare planning and National Tariff reimbursement. Data on admissions are considered very complete although there is a lag of a few weeks. Therefore, the testing data were restricted to 3 weeks prior to the date the data were linked to SUS. Hospital inpatient admissions were linked using NHS number and date of birth. Length of stay was calculated as date of discharge minus the date of admission. Admissions were restricted to those with an ICD-10 coded acute respiratory illness (ARI) discharge diagnosis in the primary diagnosis field (Supplementary Table 1), who were admitted for at least 2 days, where the date of test was 1 day before and up to 2 days after the admission (13).

### Covariates and adjustment

Vaccination status was the primary exposure variable of interest in all analyses. Additionally, all analyses included adjustment for week of test date, gender, age (five-year age bands), NHS region, IMD quintile, ethnicity and clinical risk group status (encoded as a categorial variable with a level for all conditions other than severe immunosuppression and a level for severe immunosuppression (as defined in the Green book (18)). Additionally, the analysis of long-term effectiveness in those aged 18 to 49 years included adjustment for health and social care worker status. All analyses of those aged 50 years and older included adjustment for influenza vaccination status 2023/2024. All analyses of those aged 65 years and older included adjustment for care home resident status. Less than 8% of data were missing in any category. Missing ethnicity data were included as a separate category while missing data for gender and IMD were omitted (<3% were missing).

### Statistical methods

Multivariable logistic regression was used with the test result as the outcome, vaccination status as the primary exposure variable of interest and with confounder adjustment as described above. VE was calculated as 1-odds ratio and given as a percentage. To compare estimates, statistical significance was concluded where 95% confidence intervals (CIs) did not overlap.

The residual VE (after ≥ 18 months) of two or three doses of monovalent vaccine was estimated amongst those aged 18 to 49 (as of 31^st^ August 2022 - not eligible for further boosters) who received no further doses, relative to those who were totally unvaccinated. For all other analyses except one sensitivity analysis, we estimated the incremental VE relative to those who had received at least two previous doses as we consider it most relevant to estimate the additional benefit of a booster in addition to the protection an individual has from past doses which have waned in effectiveness. The two previous doses also had to be at least 3 months prior to the test date. The residual incremental VE (after ≥ 8 months) of the bivalent BA.1 boosters given in autumn 2022 for those who got no further doses was estimated separately for those aged 50-63 years as of August 2023 (not eligible for an autumn 2023 dose) and those aged 65 years and older (eligible for an autumn 2023 dose). The residual incremental VE (in intervals 10-14, 15-19,20-25, ≥ 25 weeks post vaccination) of the spring 2023 boosters was estimated for those aged 75 years and older who had received no further doses by their test date. For this spring 2023 analysis the increment was within those who also received an autumn 2022 dose.

The incremental VE of the bivalent BA.4-5 and monovalent XBB.1.5 boosters given as part of the autumn 2023 programme was estimated for those aged 65 years and older whose last dose was administered at least 3 months prior at the following intervals since booster vaccination; 0-2 days, 3-8 days, 9-13 days, 2-4 weeks, 5-9 weeks, 10-14 weeks, and 15 or more weeks. Analyses were stratified separately by vaccine type and well as with vaccines combined. Within those cases with sequencing information available, VE was assessed by variant with manufacturers combined.

Additional analyses of the autumn 2023 boosters were conducted restricting to those aged 75 years and older, and in those aged 18 to 64 years who were in a clinical risk group. Sensitivity analyses were undertaken to assess the effect of comparing those boosted to those not boosted irrespective of prior vaccination, including adjustment for prior infection, excluding adjustment for influenza vaccine status this season (2023/2024), estimating VE against all hospitalisations, regardless of ICD-10 coding.

## Results

During the study period there were 3,428 eligible tests included in the analysis to estimate residual monovalent VE in those aged 18–49, 3,621 eligible tests included to estimate the residual autumn 2022 booster VE in 50-63 year olds, 8,790 eligible tests included to estimate the residual autumn 2022 booster VE in those aged 65 years and older, 7,123 eligible tests included to estimate the residual spring 2023 VE in those aged 75 years and older, and 28,916 eligible tests included to estimate the VE of the autumn 2023 boosters in those aged 65 years and older. Supplementary Tables 1-5 give details of the numbers of cases (positive tests) and controls (negative tests) by demographic factors and Supplementary Figure 1 details the numbers of cases and controls by calendar week.

### Residual protection of prior doses against hospitalisation

Residual incremental VE of prior doses is shown in Table 1. We found no evidence of residual VE for the monovalent boosters or the 2022 bivalent autumn boosters. The spring booster given in 2023 to those aged 75 years and over showed effectiveness waning from 30.8% (95% CI: 8.2 to 47.9%) at 10-14 weeks to no incremental protection after 20 or more weeks.

**Table 1.** Long-term duration of protection of doses given prior to the autumn 2023 vaccination programme.

| Age (years) | Vaccine | Waned period | Controls | Cases | VE (95% CI) |
| --- | --- | --- | --- | --- | --- |
| 18-49 | Unvaccinated | - | 777 | 129 | Baseline |
|  | 2 doses of monovalent vaccine | 18+ months | 826 | 134 | 1.3 (-31.2 to 25.7) |
|  | 3 doses of monovalent vaccine | 18+ months | 1378 | 184 | 9.6 (-18.4 to 31.1) |
| 50-63 | No bivalent BA.1 booster* | - | 1678 | 350 | Baseline |
|  | Bivalent BA.1 booster autumn 2022* | 8+ months | 1275 | 318 | -3.7 (-24.8 to 13.7) |
| 65+ | No bivalent BA.1 booster* | - | 2972 | 1170 | Baseline |
|  | Bivalent BA.1 booster autumn 2022* | 8+ months | 3299 | 1349 | 8.6 (-0.9 to 17.3) |
| 75+ | No spring booster** | - | 1,727 | 782 | Baseline |
|  | Spring booster 2023** | 10-14 weeks | 205 | 85 | 30.8 (8.2 to 47.9) |
|  |  | 15-19 weeks | 883 | 446 | 14.4 (-0.2 to 26.9) |
|  |  | 20-24 weeks | 1211 | 680 | -3.2 (-18.5 to 10.1) |
|  |  | 25+ weeks | 813 | 291 | -3.1 (-22.7 to 13.4) |

\* In addition to at least two prior monovalent doses, waned for at least 3 months
\*\* In addition to at least two prior monovalent doses where the last dose was a BA.1 bivalent dose given as part of the autumn 2022 programme, waned for at least 3 months

### Effectiveness of the autumn 2023 XBB.1.5 and BA.4-5 boosters against hospitalisation

The autumn 2023 booster VE peaked at 50.6% (95% CI: 44.2 to 56.3%) 2-4 weeks after vaccination, followed by waning to 13.6% (95% CI: -11.7 to 33.2%) by 15+ weeks (Table 2). The apparent VE within 0-2 days of 65.9% (95% CI: 52.5 to 75.5%) is not a true protective effect and likely a bias due to deferral of vaccination in individuals who were already aware of their COVID-19 positive status from home lateral flow testing and/or a healthy vaccinee effect in which those with early symptoms of COVID-19 were more unwell than controls and were less likely to be vaccinated. This bias should be transient as indicated by apparent VE reducing at 3-8 days to 31.0% (95% CI: 17.1 to 42.5%).

**Table 2.**
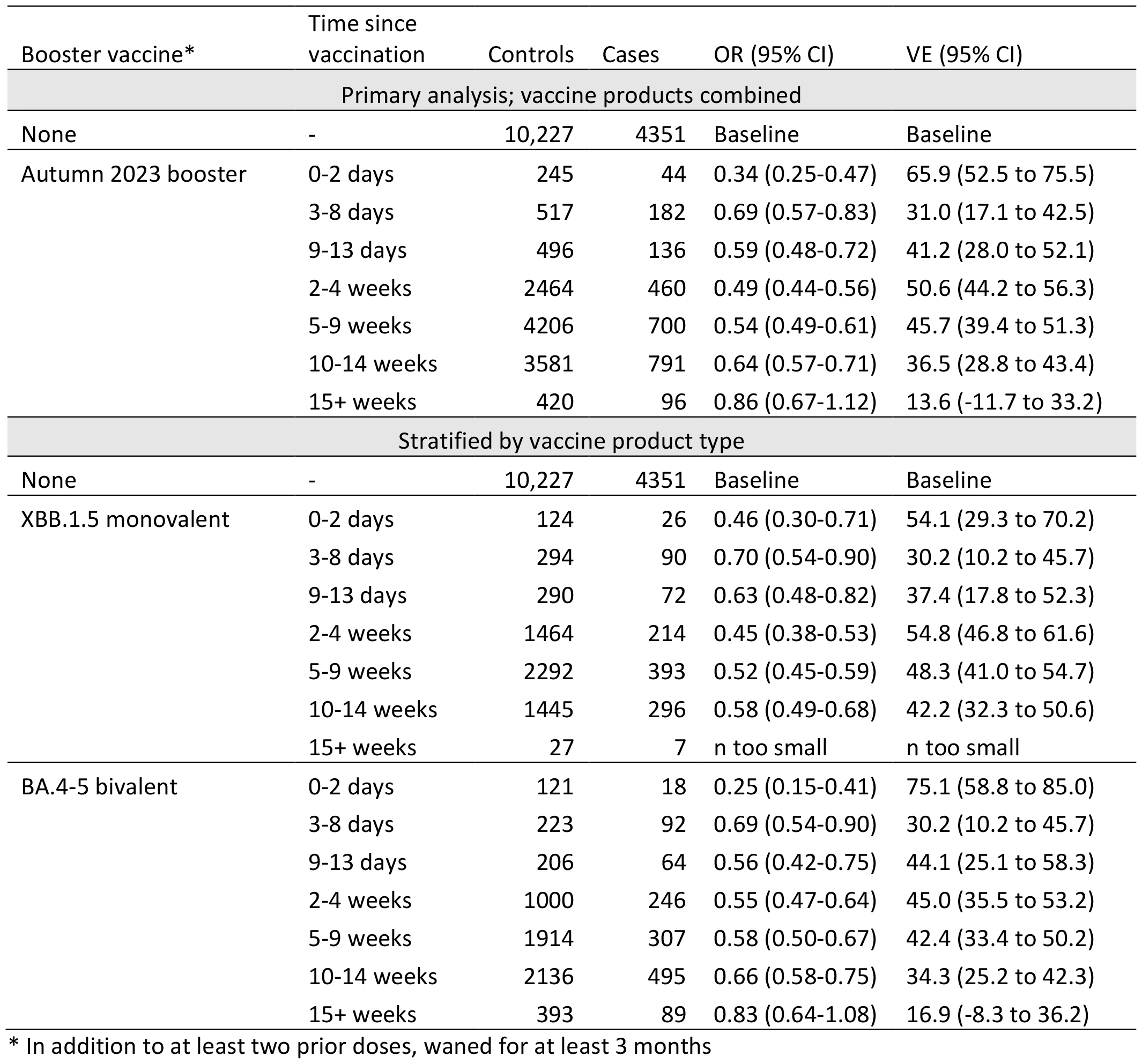
Vaccine effectiveness of the monovalent XBB.1.5 vaccine and the bivalent BA.4-5 vaccine against hospitalisation amongst those aged 65 years and older in England.

VE point estimates were generally higher for the XBB.1.5 booster as compared to the BA.4-5 booster, but confidence intervals overlapped at all time intervals investigated and this difference was therefore not statistically significant (Table 2). There were insufficient data to estimate VE at 15 or more weeks post vaccination for the XBB.1.5 booster, but VE after this period was 16.9% (95%CI: - 8.3 to 36.2%) for the BA.4-5 booster (Table 2).

When the analysis was stratified to just those aged 75 years and older, VE estimates remained similar (Supplementary Table 6). We investigated VE amongst adults aged 18 to 64 years who were in a clinical risk group and therefore also eligible for vaccination during the autumn 2023 programme (Supplementary Table 6, full descriptive characteristics in Supplementary Table 7). As the numbers of cases and controls were small, we could not stratify by vaccine type. Nonetheless, protection against hospitalisation was also observed in this population; VE was 41.5% (95%CI: 16.8 to 58.9%) after 2-4 weeks post vaccination.

Sensitivity analyses are presented in Supplementary Table 6. We conducted a sensitivity analysis where we included adjustment for past infection; this did not impact our VE estimates. We also removed the adjustment for influenza vaccination status this season (2023/2024) and this also did not alter the VE estimates. In a further sensitivity analysis, we estimated VE against all hospital admissions, regardless of the ICD-10 coding in the primary diagnosis field; this also had no significant impact on VE estimates. Finally, we assessed VE when comparing those boosted to those not boosted (irrespective of past vaccination) and found VE was also similar (Supplementary Table 6).

### Effectiveness of the autumn 2023 booster against hospitalisation with JN.1, EG.5.1 and XBB sub-lineages

For cases where sequencing was done the effectiveness of the autumn 2023 boosters against hospitalisation amongst those aged 65 years and older was estimated stratified by the Omicron sub-lineages prevalent during the study period; XBB sub-lineages, EG.5.1 and JN.1. The numbers of variant cases by calendar week in the study period are shown in Supplementary Figure 2.

Vaccine effectiveness point estimates were highest against XBB sub-lineages with VE as high as 74.0% (95%CI: 62.4 to 82.1%) at 2-4 weeks and 68.2% (95%CI: 52.9 to 78.5%) at 5-9 weeks (Table 3). VE was lower against both JN.1 and EG.5.1 with confidence intervals non-overlapping with the VE of the XBB sub-lineages at 2-4 weeks for EG.5.1 where VE was 44.5% (95% CI: 20.2 to 61.4%) and at 5-9 weeks for JN.1 where VE was 26.4% (95%CI: -3.4 to 47.6%).

**Table 3.** Vaccine effectiveness of the autumn 2023 booster vaccines against hospitalisation with JN.1, EG.5.1 and XBB sub-lineages, amongst those aged 65 years and older in England.

| Booster vaccine | Time since vaccination | Controls | Cases | OR (95% CI) | VE (95% CI) |
| --- | --- | --- | --- | --- | --- |
| JN.1 |  |  |  |  |  |
| None | - | 10,227 | 118 | Baseline | Baseline |
| Autumn 2023 booster* | 2-4 weeks | 2464 | 12 | 0.63 (0.34-1.19) | 36.6 (-19.5 to 66.4) |
|  | 5-9 weeks | 4206 | 73 | 0.74 (0.52-1.03) | 26.4 (-3.4 to 47.6) |
|  | 10+ weeks | 4001 | 163 | 0.77 (0.58-1.04) | 22.5 (-4.4 to 42.5) |
| EG.5.1 |  |  |  |  |  |
| None | - | 10,227 | 357 | Baseline | Baseline |
| Autumn 2023 booster* | 2-4 weeks | 2464 | 48 | 0.55 (0.39-0.80) | 44.5 (20.2 to 61.4) |
|  | 5-9 weeks | 4206 | 40 | 0.57 (0.37-0.86) | 43.2 (13.7 to 62.7) |
|  | 10+ weeks | 4001 | 21 | 0.78 (0.43-1.43) | 21.6 (-43.0 to 57.0) |
| XBB sub-lineages |  |  |  |  |  |
| None | - | 10,227 | 649 | Baseline | Baseline |
| Autumn 2023 booster* | 2-4 weeks | 2464 | 37 | 0.26 (0.18-0.38) | 74.0 (62.4 to 82.1) |
|  | 5-9 weeks | 4206 | 37 | 0.32 (0.22-0.47) | 68.2 (52.9 to 78.5) |
|  | 10+ weeks | 4001 | 22 | 0.45 (0.26-0.77) | 55.1 (23.1 to 73.8) |

\*Of the bivalent BA.4-5 booster or the monovalent XBB.1.5 booster

After 10 or more weeks, the protection against hospitalisation was 22.5% (95%CI: -4.4 to 42.5%), 21.6% (95%CI: -43.0 to 57.0%) and 55.1% (95% CI: 23.1 to 73.8%) for JN.1, EG.5.1 and XBB sub-lineages, respectively (Table 3). It should be noted that in this longer period post vaccination numbers are small for the JN.1 and EG.5.1 analyses, and the mean time since vaccination within this interval will be shorter than that for JN.1 (the distribution of time since vaccination by variant is shown in Supplementary Figure 3).

## Conclusions

We here used a TNCC study design to estimate protection from the COVID-19 vaccines against hospitalisation amongst all adults in England, depending on which vaccine individuals were last eligible for. Overall, we find that effectiveness of doses given prior to the most recent autumn 2023 booster now provide minimal protection against hospitalisation. The most recent XBB.1.5 and BA.4-5 boosters given as part of the autumn 2023 programme provided good protection amongst those aged 65 years and older. Protection peaked at around 40-50% against hospitalisation in addition to the waned immunity from prior doses. There was evidence of waning of protection of these boosters, but this was partially confounded by the emergence of the JN.1 variant in England towards the end of the study period for which we found evidence of reduced VE.

Adults aged 18 to 49 years in England were last offered a two-dose primary course and booster (third dose) from November 2021. The current effectiveness of these doses after at least 18 months was minimal; there was no longer any significant protection against hospitalisation as compared to their unvaccinated peers. Previously, we observed high VE against hospitalisation for these vaccines, but low VE was observed against mild disease (13, 14, 19) and infection (20). Since, there have been multiple waves of infections and therefore many individuals’ (both vaccinated and unvaccinated) most recent immunising event will be an infection. The reduced protection we observed is therefore partly attributable to waning but also likely reflective of the very high level of immunity in both unvaccinated and vaccinated individuals from waves of prior infections over the last two years. COVID-19 hospitalisation rates remain very low for younger adults and England’s national surveillance data do not indicate that this population is at risk of severe disease (5).

The incremental VE of the BA.1 bivalent booster offered in the autumn of 2022 and of the spring booster administered in 2023 was also found to be minimal after 8 months and after 5 months, respectively, indicating that these doses are no longer providing additional protection against hospitalisation as compared to those who had received at least two doses but had not received a booster as part of that programme. This is also likely also due to a combination of waning and recent infections increasing the immunity in the comparator group to a comparable level to those vaccinated. Although waves of COVID-19 continue to occur, the magnitude of hospitalisations has decreased with each wave (21), indicating that those waned immunity are not necessarily at increased risk of severe disease. Policy decisions around the timing of vaccination must consider the short-lived duration of protection to maximise protection at the right time for those at higher risk of severe disease. This may be challenging as COVID-19 waves do not follow a seasonal pattern and occur throughout the year due to the emergence of novel sub-lineages, the timing of which is difficult to predict.

The effectiveness of the most recent boosters, the monovalent XBB.1.5 and bivalent BA.4-5 vaccines, amongst those aged 65 years was comparable to the incremental protection we observed previously shortly after vaccination with the bivalent BA.1 boosters (1) and with the Sanofi/GSK product (2). We did not observe a significant difference in protection by vaccine type. Early data using a cohort study design from Denmark found higher VE of the XBB.1.5 boosters - in their study vaccination was associated with a 76% reduced risk of COVID-19 hospitalisation (22). However, the average follow-up time post vaccination was just 9.9 days. VE results from TNCC studies from the US and Canada were broadly similar to our study and found VE of the XBB.1.5 boosters to be 46% against symptomatic infection (23) and 47% against medically attended outpatient COVID-19 (24). In a stratified analysis we found VE of was similar for those aged 75 years and older who are considered at higher risk of severe disease. We also found moderate protection in adults aged 18-64 years who were in a clinical risk group and therefore also eligible for vaccination. Point estimates for younger adults in a clinical risk group were lower than the VE estimates for adults aged 65 years and older, although numbers of cases and controls included in the analysis were smaller and therefore the precision around the estimates was also lower. We found VE was similar if we simply compared those boosted to those not boosted, rather than restricting to the analysis being within those with at least 2 prior doses. This is not surprising given the minimal residual VE of prior doses seen.

Since freely available community COVID-19 testing ended in April 2021 in England, most infections are now unreported. In sensitivity analyses we found that there was no difference between the VE estimates when we included adjustment for the likely variant of a person’s most recent prior infection in the model. This is in agreement with previous studies where we have also not found a difference (1, 2). Influenza vaccination status this season (2023/2024) was adjusted for in the primary analysis. An assumption of the TNCC design is that the risk of being a control (a non-COVID-19 respiratory hospitalisation) is independent of vaccination status. This may not be the case when influenza and COVID-19 are co-circulating as those receiving a COVID-19 vaccine are also more likely to receive an influenza vaccine (25). One option, if all samples were tested for influenza would be to remove these controls, but as this was not done we instead adjusted for influenza vaccination status despite a level of collinearity with COVID-19 vaccination. Reassuringly, we did not find that influenza vaccination had a protective effect against COVID-19 hospitalisation and in sensitivity analyses we did not find that removing the adjustment for influenza vaccination status had an effect on the VE estimates.

There was evidence of waning after 10 weeks post vaccination for both the XBB.1.5 and BA.4-5 boosters. This is also in line with when we have observed waning for previous boosters (1, 2) but this was partially confounded by the emergence of JN.1 during the study period. A study of vaccine effectiveness (VE) against symptomatic disease from the US found no significant difference in VE between likely JN.1 and non-JN.1 cases (identified by S-gene target status) (23) however a study from Denmark found that JN.1 cases had higher odds of being vaccinated than non-BA.2.86 cases – suggesting lower real-world effectiveness (26). In agreement with the Danish study, we also found evidence of reduced effectiveness in the period 5-9 weeks post-vaccination. This difference was significant when comparing VE against JN.1 and XBB-related sub-lineages but not significant when comparing VE against JN.1 and EG.5.1, possibly due to the XBB.1.5 vaccine being better matched to XBB-related sub-lineages. While England saw a wave of cases driven by the emergence of JN.1, which peaked around week 01 of 2024, this has since declined and was similar or smaller in magnitude to previous waves (5), suggesting that despite lower VE, population-wide immunity (from vaccines or natural infections) has suppressed the JN.1 wave in England.

A strength of this study is the use of national-level electronic healthcare datasets to estimate VE against hospitalisation for the population of England. To our knowledge, this is the only recent study which has investigated the long-term duration of protection in the whole population, therefore giving insight into the status of the population as a whole and not just those offered the most recent booster. A key strength is also the use of the TNCC study design which helps to address unmeasured confounders related to differences in health-seeking behaviours, health care access and infectious-disease exposure between vaccinated and unvaccinated people. Nonetheless our study has several limitations; even with the use of national-level datasets, COVID-19 testing and whole genome sequencing has been scaled down which limits the speed at which we able to estimate VE in response to emergent variants, and the precision around the estimates. It is therefore more challenging to tease apart smaller differences in VE between vaccine types and co-circulating variants. Additionally, since this is an observational study, there may have been unmeasured or residual confounding which we were not able to adjust for.

Overall, we here find evidence that those vaccinated during earlier campaigns in England are no longer better protected than their unvaccinated counterparts. The recent monovalent XBB.1.5 and bivalent BA.4-5 boosters provided comparable and good protection against hospitalisation amongst those aged 65 years and older, however there was evidence of lower VE against hospitalisation of these boosters against JN.1.

## Data Availability

This work is carried out under Regulation 3 of The Health Service (Control of Patient Information; Secretary of State for Health, 2002) using patient identification information without individual patient consent as part of the UKHSA legal requirement for public health surveillance and monitoring of vaccines. As such, authors cannot make the underlying dataset publicly available for ethical and legal reasons. However, all the data used for this analysis is included as aggregated data in the manuscript tables and appendix. Applications for relevant anonymised data should be submitted to the UKHSA Office for Data Release at https://www.gov.uk/government/publications/accessing-ukhsa-protected-data.

## Authors’ Contributions

NA, FCMK and JLB conceptualised the study. FCMK, JS and NA developed the methodology. FCMK curated the data. FCMK accessed and verified the data. FCMK conducted the formal analysis, supported by NA. FCMK wrote the original draft of the manuscript. JLB and AA provided supervision. All co-authors reviewed the manuscript and were responsible for the decision to submit the manuscript.

### Ethics Committee Approval

The study protocol was subject to an internal review by the UK Health Security Agency Research Ethics and Governance Group and was found to be fully compliant with all regulatory requirements. As no regulatory issues were identified, and ethical review is not a requirement for this type of work, it was decided that a full ethical review would not be necessary. UKHSA has legal permission, provided by Regulation 3 of The Health Service (Control of Patient Information) Regulations 2002, to process patient confidential information for national surveillance of communicable diseases and as such, individual patient consent is not required to access records.

## References

1. Kirsebom FCM, Andrews N, Stowe J, Ramsay M, Lopez Bernal J. Duration of protection of ancestral-strain monovalent vaccines and effectiveness of bivalent BA.1 boosters against COVID-19 hospitalisation in England: a test-negative case-control study. The Lancet Infectious Diseases. 2023;0(0).

2. Kirsebom FCM, Andrews N, Stowe J, Dabrera G, Ramsay M, Bernal JL. Effectiveness of the adjuvanted Sanofi/GSK (VidPrevtyn Beta) and Pfizer-BioNTech (Comirnaty Original/Omicron BA.4-5) bivalent vaccines against hospitalisation amongst adults aged 75 years and older in England, estimated using a test-negative case control study design. medRxiv. 2023:2023.09.28.23296290.

3. Link-Gelles R, Ciesla AA, Roper LE, Scobie HM, Ali AR, Miller JD, et al. Early Estimates of Bivalent mRNA Booster Dose Vaccine Effectiveness in Preventing Symptomatic SARS-CoV-2 Infection Attributable to Omicron BA.5- and XBB/XBB.1.5-Related Sublineages Among Immunocompetent Adults - Increasing Community Access to Testing Program, United States, December 2022-January 2023. MMWR Morbidity and mortality weekly report. 2023;72(5):119–24.

4. Tenforde Mw Fau - Weber ZA, Weber Za Fau - Natarajan K, Natarajan K Fau - Klein NP, Klein Np Fau - Kharbanda AB, Kharbanda Ab Fau - Stenehjem E, Stenehjem E Fau - Embi PJ, et al. Early Estimates of Bivalent mRNA Vaccine Effectiveness in Preventing COVID-19-Associated Emergency Department or Urgent Care Encounters and Hospitalizations Among Immunocompetent Adults - VISION Network, Nine States, September-November 2022. MMWR Morbidity and mortality weekly report. 2022(1545-861X (Electronic)).

5. UK Health Security Agency. National Influenza and COVID-19 surveillance report: Week 9 report (up to week 8 data) 2024 [Available from: https://assets.publishing.service.gov.uk/media/65e07c292f2b3b001c7cd778/Weekly-flu-and-COVID-19-surveillance-report-week-9.pdf.

6. Yang S, Yu Y, Xu Y, Jian F, Song W, Yisimayi A, et al. Fast evolution of SARS-CoV-2 BA.2.86 to JN.1 under heavy immune pressure. The Lancet Infectious diseases. 2024;24(2):e70–e2.

7. World Health Organisation. Initial risk evaluation of JN.1, 19 December 2023 2023 [Available from: https://www.who.int/docs/default-source/coronaviruse/18122023_jn.1_ire_clean.pdf?sfvrsn=6103754a_3.

8. Department of Health & Social Care. Joint Committee on Vaccination and Immunisation: advice on priority groups for COVID-19 vaccination, 30 December 2020 [Available from: https://www.gov.uk/government/publications/priority-groups-for-coronavirus-covid-19-vaccination-advice-from-the-jcvi-30-december-2020/joint-committee-on-vaccination-and-immunisation-advice-on-priority-groups-for-covid-19-vaccination-30-december-2020.

9. Joint Committee on Vaccination and Immunisation. Joint Committee on Vaccination and Immunisation (JCVI) statement on COVID-19 vaccinations in 2022: 21 February 2022. 2022.

10. Joint Committee on Vaccination and Immunisation. JCVI statement on spring 2023 COVID-19 vaccinations, 22 February 2023 2023 [Available from: https://www.gov.uk/government/publications/spring-2023-covid-19-vaccination-programme-jcvi-advice-22-february-2023/jcvi-statement-on-spring-2023-covid-19-vaccinations-22-february-2023.

11. Joint Committee on Vaccination and Immunisation. JCVI updated statement on the COVID-19 vaccination programme for autumn 2022 2022 [Available from: https://www.gov.uk/government/publications/jcvi-updated-statement-on-the-covid-19-vaccination-programme-for-autumn-2022.

12. Joint Committee on Vaccination and Immunisation. JCVI statement on the COVID-19 vaccination programme for autumn 2023 - update 7 July 2023 2024 [Available from: https://www.gov.uk/government/publications/covid-19-autumn-2023-vaccination-programme-jcvi-update-7-july-2023/jcvi-statement-on-the-covid-19-vaccination-programme-for-autumn-2023-update-7-july-2023.

13. Stowe J, Andrews N, Kirsebom F, Ramsay M, Bernal JL. Effectiveness of COVID-19 vaccines against Omicron and Delta hospitalisation, a test negative case-control study. Nature communications. 2022;13(1):5736.

14. Andrews N, Stowe J, Kirsebom F, Toffa S, Rickeard T, Gallagher E, et al. Covid-19 Vaccine Effectiveness against the Omicron (B.1.1.529) Variant. New England Journal of Medicine. 2022.

15. Tessier E, Edelstein M, Tsang C, Kirsebom F, Gower C, Campbell CNJ, et al. Monitoring the COVID-19 immunisation programme through a national immunisation Management system – England’s experience. International Journal of Medical Informatics. 2023;170:104974.

16. NHS Digital. Cohorting as a Service (CaaS) 2022 [Available from: https://digital.nhs.uk/services/cohorting-as-a-service-caas.

17. NHS Digital. Secondary Uses Service (SUS) 2022 [Available from: https://digital.nhs.uk/services/secondary-uses-service-sus.

18. UK Health Security Agency. COVID-19: the green book, chapter 14a. Immunisation against infectious diseases: UK Health Security Agency,; 2020.

19. Kirsebom FCM, Andrews N, Stowe J, Toffa S, Sachdeva R, Gallagher E, et al. COVID-19 vaccine effectiveness against the omicron (BA.2) variant in England. The Lancet Infectious Diseases. 2022;22(7):931–3.

20. Kirwan PD, Hall VJ, Foulkes S, Otter AD, Munro K, Sparkes D, et al. Effect of second booster vaccinations and prior infection against SARS-CoV-2 in the UK SIREN healthcare worker cohort. Lancet Reg Health Eur. 2024;36:100809.

21. UK Health Security Agency. UK Coronavirus Dashboard; Healthcare in England 2024 [Available from: https://coronavirus.data.gov.uk/details/healthcare?areaType=nation&areaName=England.

22. Hansen CH, Moustsen-Helms IR, Rasmussen M, Soborg B, Ullum H, Valentiner-Branth P. Short-term effectiveness of the XBB.1.5 updated COVID-19 vaccine against hospitalisation in Denmark: a national cohort study. The Lancet Infectious diseases. 2024;24(2):e73–e4.

23. Link-Gelles R, Ciesla AA, Mak J, Miller JD, Silk BJ, Lambrou AS, et al. Early Estimates of Updated 2023-2024 (Monovalent XBB.1.5) COVID-19 Vaccine Effectiveness Against Symptomatic SARS-CoV-2 Infection Attributable to Co-Circulating Omicron Variants Among Immunocompetent Adults - Increasing Community Access to Testing Program, United States, September 2023-January 2024. MMWR Morbidity and mortality weekly report. 2024;73(4):77–83.

24. Skowronski DM, Zhan Y, Kaweski SE, Sabaiduc S, Khalid A, Olsha R, et al. 2023/24 mid-season influenza and Omicron XBB.1.5 vaccine effectiveness estimates from the Canadian Sentinel Practitioner Surveillance Network (SPSN). Euro surveillance : bulletin Europeen sur les maladies transmissibles = European communicable disease bulletin. 2024;29(7).

25. Doll MK, Pettigrew SM, Ma J, Verma A. Effects of Confounding Bias in Coronavirus Disease 2019 (COVID-19) and Influenza Vaccine Effectiveness Test-Negative Designs Due to Correlated Influenza and COVID-19 Vaccination Behaviors. Clinical infectious diseases : an official publication of the Infectious Diseases Society of America. 2022;75(1):e564–e71.

26. Moustsen-Helms I, Bager P, Larsen T, Trier Møller F, Vestergaard L, Christiansen L, et al. Relative Vaccine Protection, Disease Severity and Symptoms Associated with Infection with SARS-CoV-2 Omicron Subvariant Ba.2.86 and Descendent Jn.1: A Danish Nationwide Register-Based Study. Preprint with SSRN. 2024.

