## Supplementary Appendix for "Effectiveness of Autumn 2023 COVID-19 vaccination and residual protection of prior doses against hospitalisation in England, estimated using a test-negative case-control study"

Supplementary Table 1. Descriptive characteristics of cases and controls included in the analysis of the effectiveness of the monovalent vaccines against hospitalisation amongst those aged 18 to 49 years, as presented in Table 1. Cases and controls by calendar week are shown in Supplementary Figure 1.

| | | Overall | | Controls | | Cases | |
|---|---|---|---|---|---|---|---|
| | | n | % | n | % | n | % |
| | Totals | 3428 | 100% | 2,981 | 87% | 447 | 13% |
| Gender | Female | 1,860 | 54.3% | 1,563 | 52.4% | 297 | 66.4% |
| | Male | 1,487 | 43.4% | 1,337 | 44.9% | 150 | 33.6% |
| | Missing | 81 | 2.4% | 81 | 2.7% | 0 | 0.0% |
| Age | 18-19 | 123 | 3.6% | 101 | 3.4% | 22 | 4.9% |
| | 20-24 | 324 | 9.5% | 264 | 8.9% | 60 | 13.4% |
| | 25-29 | 489 | 14.3% | 421 | 14.1% | 68 | 15.2% |
| | 30-34 | 571 | 16.7% | 504 | 16.9% | 67 | 15.0% |
| | 35-39 | 616 | 18.0% | 550 | 18.5% | 66 | 14.8% |
| | 40-44 | 658 | 19.2% | 579 | 19.4% | 79 | 17.7% |
| | 45-49 | 647 | 18.9% | 562 | 18.9% | 85 | 19.0% |
| NHS Region | East of England | 245 | 7.1% | 219 | 7.3% | 26 | 5.8% |
| | London | 600 | 17.5% | 511 | 17.1% | 89 | 19.9% |
| | Midlands | 830 | 24.2% | 694 | 23.3% | 136 | 30.4% |
| | North East | 600 | 17.5% | 529 | 17.7% | 71 | 15.9% |
| | North West | 513 | 15.0% | 466 | 15.6% | 47 | 10.5% |
| | South East | 397 | 11.6% | 350 | 11.7% | 47 | 10.5% |
| | South West | 243 | 7.1% | 212 | 7.1% | 31 | 6.9% |
| Ethnicity | African | 98 | 2.9% | 77 | 2.6% | 21 | 4.7% |
| | Any other Asian background | 91 | 2.7% | 70 | 2.3% | 21 | 4.7% |
| | Any other Black background | 48 | 1.4% | 42 | 1.4% | 6 | 1.3% |
| | Any other White background | 299 | 8.7% | 257 | 8.6% | 42 | 9.4% |
| | Any other ethnic group | 111 | 3.2% | 90 | 3.0% | 21 | 4.7% |
| | Any other mixed background | 32 | 0.9% | 28 | 0.9% | 4 | 0.9% |
| | Bangladeshi or British Bangladeshi | 59 | 1.7% | 52 | 1.7% | 7 | 1.6% |
| | British, Mixed British | 2,018 | 58.9% | 1,786 | 59.9% | 232 | 51.9% |
| | Caribbean | 46 | 1.3% | 34 | 1.1% | 12 | 2.7% |
| | Chinese | 7 | 0.2% | 7 | 0.2% | 0 | 0.0% |
| | Indian or British Indian | 131 | 3.8% | 113 | 3.8% | 18 | 4.0% |
| | Irish | 17 | 0.5% | 16 | 0.5% | 1 | 0.2% |
| | Pakistani or British Pakistani | 167 | 4.9% | 138 | 4.6% | 29 | 6.5% |
| | White and Asian | 17 | 0.5% | 17 | 0.6% | 0 | 0.0% |
| | White and Black African | 12 | 0.4% | 9 | 0.3% | 3 | 0.7% |
| | White and Black Caribbean | 22 | 0.6% | 16 | 0.5% | 6 | 1.3% |
| | Missing | 253 | 7.4% | 229 | 7.7% | 24 | 5.4% |
| IMD Quintiles | 1 | 1,091 | 31.8% | 940 | 31.5% | 151 | 33.8% |
| | 2 | 785 | 22.9% | 671 | 22.5% | 114 | 25.5% |
| | 3 | 620 | 18.1% | 540 | 18.1% | 80 | 17.9% |

| | | | | | | | |
|---|---|---|---|---|---|---|---|
| | 4 | 470 | 13.7% | 408 | 13.7% | 62 | 13.9% |
| | 5 | 414 | 12.1% | 380 | 12.7% | 34 | 7.6% |
| | Missing | 48 | 1.4% | 42 | 1.4% | 6 | 1.3% |
| Risk Status | Health & social care worker | 147 | 4.3% | 128 | 4.3% | 19 | 4.3% |
| | Clinical risk group (excl. severely immunosuppressed) | 1,297 | 37.8% | 1,090 | 36.6% | 207 | 46.3% |
| | Severely immunosuppressed | 144 | 4.2% | 111 | 3.7% | 33 | 7.4% |
| COVID vaccine status | Unvaccinated | 906 | 26% | 777 | 26% | 129 | 29% |
| | Two doses (monovalent), waned for 18+ months | 960 | 28.0% | 826 | 27.7% | 134 | 30.0% |
| | Three doses (monovalent), waned for 18+ months | 1,562 | 45.6% | 1,378 | 46.2% | 184 | 41.2% |

Supplementary Table 2. Descriptive characteristics of cases and controls included in the analysis of the effectiveness of bivalent BA.1 boosters given in autumn 2022 against hospitalisation amongst those aged 50 to 63 years, as presented in Table 1. Cases and controls by calendar week are shown in Supplementary Figure 1.

| | | Overall | | Controls | | Cases | |
|---|---|---|---|---|---|---|---|
| | | n | % | n | % | n | % |
| | Totals | 3,621 | 100% | 2,953 | 82% | 668 | 18% |
| Gender | Female | 1,814 | 50.1% | 1,456 | 49.3% | 358 | 53.6% |
| | Male | 1,718 | 47.4% | 1,410 | 47.7% | 308 | 46.1% |
| | Missing | 89 | 2.5% | 87 | 2.9% | 2 | 0.3% |
| Age | 50-54 | 1,051 | 29.0% | 882 | 29.9% | 169 | 25.3% |
| | 55-59 | 1,345 | 37.1% | 1,093 | 37.0% | 252 | 37.7% |
| | 60-64 | 1,225 | 33.8% | 978 | 33.1% | 247 | 37.0% |
| NHS Region | East of England | 272 | 7.5% | 232 | 7.9% | 40 | 6.0% |
| | London | 531 | 14.7% | 443 | 15.0% | 88 | 13.2% |
| | Midlands | 868 | 24.0% | 694 | 23.5% | 174 | 26.0% |
| | North East | 672 | 18.6% | 555 | 18.8% | 117 | 17.5% |
| | North West | 584 | 16.1% | 483 | 16.4% | 101 | 15.1% |
| | South East | 421 | 11.6% | 329 | 11.1% | 92 | 13.8% |
| | South West | 273 | 7.5% | 217 | 7.3% | 56 | 8.4% |
| Ethnicity | African | 54 | 1.5% | 45 | 1.5% | 9 | 1.3% |
| | Any other Asian background | 58 | 1.6% | 49 | 1.7% | 9 | 1.3% |
| | Any other Black background | 25 | 0.7% | 24 | 0.8% | 1 | 0.1% |
| | Any other White background | 194 | 5.4% | 159 | 5.4% | 35 | 5.2% |
| | Any other ethnic group | 64 | 1.8% | 55 | 1.9% | 9 | 1.3% |
| | Any other mixed background | 20 | 0.6% | 15 | 0.5% | 5 | 0.7% |
| | Bangladeshi or British Bangladeshi | 32 | 0.9% | 31 | 1.0% | 1 | 0.1% |
| | British, Mixed British | 2,738 | 75.6% | 2,209 | 74.8% | 529 | 79.2% |
| | Caribbean | 50 | 1.4% | 38 | 1.3% | 12 | 1.8% |
| | Chinese | 10 | 0.3% | 10 | 0.3% | 0 | 0.0% |
| | Indian or British Indian | 109 | 3.0% | 91 | 3.1% | 18 | 2.7% |
| | Irish | 33 | 0.9% | 29 | 1.0% | 4 | 0.6% |
| | Pakistani or British Pakistani | 101 | 2.8% | 88 | 3.0% | 13 | 1.9% |
| | White and Asian | 5 | 0.1% | 4 | 0.1% | 1 | 0.1% |
| | White and Black African | 5 | 0.1% | 3 | 0.1% | 2 | 0.3% |
| | White and Black Caribbean | 7 | 0.2% | 6 | 0.2% | 1 | 0.1% |
| | Missing | 116 | 3.2% | 97 | 3.3% | 19 | 2.8% |
| IMD Quintiles | 1 | 1,127 | 31.1% | 950 | 32.2% | 177 | 26.5% |
| | 2 | 804 | 22.2% | 651 | 22.0% | 153 | 22.9% |
| | 3 | 655 | 18.1% | 519 | 17.6% | 136 | 20.4% |
| | 4 | 589 | 16.3% | 480 | 16.3% | 109 | 16.3% |
| | 5 | 420 | 11.6% | 333 | 11.3% | 87 | 13.0% |
| | Missing | 26 | 0.7% | 20 | 0.7% | 6 | 0.9% |
| Risk Status | Health & social care worker | 108 | 3.0% | 91 | 3.1% | 17 | 2.5% |

| | | | | | | | |
|---|---|---|---|---|---|---|---|
| | Clinical risk group (excl. severely immunosuppressed) | 2,363 | 65.3% | 1,860 | 63.0% | 503 | 75.3% |
| | Severely immunosuppressed | 241 | 6.7% | 179 | 6.1% | 62 | 9.3% |
| Influenza vaccine 2023/2024 | | 544 | 15% | 453 | 15% | 91 | 14% |
| COVID vaccine status | Not boosted with BA.1 bivalent* | 2,028 | 56% | 1678 | 57% | 350 | 52% |
| | BA.1 bivalent booster, waned for at least 8 months* | 1,593 | 44.0% | 1,275 | 43.2% | 318 | 47.6% |

*In addition to at least two monovalent doses, waned for at least 3 months

Supplementary Table 3. Descriptive characteristics of cases and controls included in the analysis of the effectiveness of bivalent BA.1 boosters given in autumn 2022 against hospitalisation amongst those aged 65 years and older, as presented in Table 1. Cases and controls by calendar week are shown in Supplementary Figure 1.

| | | Overall | | Controls | | Cases | |
|---|---|---|---|---|---|---|---|
| | | n | % | n | % | n | % |
| | Totals | 8,790 | 100% | 6,271 | 71% | 2519 | 29% |
| Gender | Female | 4,619 | 52.5% | 3,346 | 53.4% | 1,273 | 50.5% |
| | Male | 4,042 | 46.0% | 2,798 | 44.6% | 1,244 | 49.4% |
| | Missing | 129 | 1.5% | 127 | 2.0% | 2 | 0.1% |
| Age | 65-69 | 1,677 | 19.1% | 1,278 | 20.4% | 399 | 15.8% |
| | 70-74 | 2,062 | 23.5% | 1,492 | 23.8% | 570 | 22.6% |
| | 75-79 | 1,577 | 17.9% | 1,114 | 17.8% | 463 | 18.4% |
| | 80-84 | 1,404 | 16.0% | 957 | 15.3% | 447 | 17.7% |
| | 85-89 | 1,210 | 13.8% | 827 | 13.2% | 383 | 15.2% |
| | 90+ | 860 | 9.8% | 603 | 9.6% | 257 | 10.2% |
| NHS Region | East of England | 578 | 6.6% | 399 | 6.4% | 179 | 7.1% |
| | London | 1,563 | 17.8% | 1,165 | 18.6% | 398 | 15.8% |
| | Midlands | 2,102 | 23.9% | 1,422 | 22.7% | 680 | 27.0% |
| | North East | 1,704 | 19.4% | 1,210 | 19.3% | 494 | 19.6% |
| | North West | 1,345 | 15.3% | 1,030 | 16.4% | 315 | 12.5% |
| | South East | 897 | 10.2% | 633 | 10.1% | 264 | 10.5% |
| | South West | 601 | 6.8% | 412 | 6.6% | 189 | 7.5% |
| Ethnicity | African | 73 | 0.8% | 54 | 0.9% | 19 | 0.8% |
| | Any other Asian background | 148 | 1.7% | 104 | 1.7% | 44 | 1.7% |
| | Any other Black background | 30 | 0.3% | 23 | 0.4% | 7 | 0.3% |
| | Any other White background | 457 | 5.2% | 324 | 5.2% | 133 | 5.3% |
| | Any other ethnic group | 129 | 1.5% | 97 | 1.5% | 32 | 1.3% |
| | Any other mixed background | 36 | 0.4% | 23 | 0.4% | 13 | 0.5% |
| | Bangladeshi or British Bangladeshi | 78 | 0.9% | 61 | 1.0% | 17 | 0.7% |
| | British, Mixed British | 6,672 | 75.9% | 4,690 | 74.8% | 1,982 | 78.7% |
| | Caribbean | 85 | 1.0% | 59 | 0.9% | 26 | 1.0% |
| | Chinese | 10 | 0.1% | 7 | 0.1% | 3 | 0.1% |
| | Indian or British Indian | 322 | 3.7% | 253 | 4.0% | 69 | 2.7% |
| | Irish | 136 | 1.5% | 108 | 1.7% | 28 | 1.1% |
| | Pakistani or British Pakistani | 241 | 2.7% | 196 | 3.1% | 45 | 1.8% |
| | White and Asian | 9 | 0.1% | 4 | 0.1% | 5 | 0.2% |
| | White and Black African | 8 | 0.1% | 6 | 0.1% | 2 | 0.1% |
| | White and Black Caribbean | 14 | 0.2% | 7 | 0.1% | 7 | 0.3% |
| | Missing | 342 | 3.9% | 255 | 4.1% | 87 | 3.5% |
| IMD Quintiles | 1 | 2,504 | 28.5% | 1,822 | 29.1% | 682 | 27.1% |
| | 2 | 1,975 | 22.5% | 1,412 | 22.5% | 563 | 22.4% |
| | 3 | 1,664 | 18.9% | 1,199 | 19.1% | 465 | 18.5% |
| | 4 | 1,483 | 16.9% | 1,019 | 16.2% | 464 | 18.4% |

| | | | | | | | |
|---|---|---|---|---|---|---|---|
| | 5 | 1,138 | 12.9% | 804 | 12.8% | 334 | 13.3% |
| | Missing | 26 | 0.3% | 15 | 0.2% | 11 | 0.4% |
| Risk Status | Health & social care worker | 37 | 0.4% | 27 | 0.4% | 10 | 0.4% |
| | Clinical risk group (excl. severely immunosuppressed) | 7,393 | 84.1% | 5,233 | 83.4% | 2,160 | 85.7% |
| | Severely immunosuppressed | 519 | 5.9% | 364 | 5.8% | 155 | 6.2% |
| Carehome resident | | 151 | 2% | 124 | 2% | 27 | 1% |
| Influenza vaccine 2023/2024 | | 1,841 | 21% | 1381 | 22% | 460 | 18% |
| COVID vaccine status | Not boosted with BA.1 bivalent* | 4,142 | 47% | 2,972 | 47% | 1,170 | 46% |
| | BA.1 bivalent booster, waned for at least 8 months* | 4,648 | 52.9% | 3,299 | 52.6% | 1,349 | 53.6% |

*In addition to at least two monovalent doses, waned for at least 3 months

Supplementary Table 4. Descriptive characteristics of cases and controls included in the analysis of the effectiveness of the spring 2023 boosters against hospitalisation amongst those aged 75 years and older, as presented in Table 1. Cases and controls by calendar week are shown in Supplementary Figure 1.

| | | Overall | | Controls | | Cases | |
|---|---|---|---|---|---|---|---|
| | | n | % | n | % | n | % |
| | Totals | 7,123 | 100% | 4,839 | 68% | 2,284 | 32% |
| Gender | Female | 3,601 | 50.6% | 2,507 | 51.8% | 1,094 | 47.9% |
| | Male | 3,415 | 47.9% | 2,226 | 46.0% | 1,189 | 52.1% |
| | Missing | 107 | 1.5% | 106 | 2.2% | 1 | 0.0% |
| | 75-79 | 1,970 | 27.7% | 1,372 | 28.4% | 598 | 26.2% |
| | 80-84 | 1,933 | 27.1% | 1,298 | 26.8% | 635 | 27.8% |
| | 85-89 | 1,796 | 25.2% | 1,186 | 24.5% | 610 | 26.7% |
| | 90+ | 1,424 | 20.0% | 983 | 20.3% | 441 | 19.3% |
| NHS Region | East of England | 584 | 8.2% | 396 | 8.2% | 188 | 8.2% |
| | London | 909 | 12.8% | 666 | 13.8% | 243 | 10.6% |
| | Midlands | 1,756 | 24.7% | 1,135 | 23.5% | 621 | 27.2% |
| | North East | 1,395 | 19.6% | 932 | 19.3% | 463 | 20.3% |
| | North West | 886 | 12.4% | 639 | 13.2% | 247 | 10.8% |
| | South East | 926 | 13.0% | 610 | 12.6% | 316 | 13.8% |
| | South West | 667 | 9.4% | 461 | 9.5% | 206 | 9.0% |
| Ethnicity | African | 20 | 0.3% | 15 | 0.3% | 5 | 0.2% |
| | Any other Asian background | 55 | 0.8% | 43 | 0.9% | 12 | 0.5% |
| | Any other Black background | 6 | 0.1% | 4 | 0.1% | 2 | 0.1% |
| | Any other White background | 321 | 4.5% | 219 | 4.5% | 102 | 4.5% |
| | Any other ethnic group | 58 | 0.8% | 43 | 0.9% | 15 | 0.7% |
| | Any other mixed background | 19 | 0.3% | 10 | 0.2% | 9 | 0.4% |
| | Bangladeshi or British Bangladeshi | 12 | 0.2% | 9 | 0.2% | 3 | 0.1% |
| | British, Mixed British | 6,015 | 84.4% | 4,069 | 84.1% | 1,946 | 85.2% |
| | Caribbean | 30 | 0.4% | 20 | 0.4% | 10 | 0.4% |
| | Chinese | 12 | 0.2% | 10 | 0.2% | 2 | 0.1% |
| | Indian or British Indian | 142 | 2.0% | 102 | 2.1% | 40 | 1.8% |
| | Irish | 99 | 1.4% | 60 | 1.2% | 39 | 1.7% |
| | Pakistani or British Pakistani | 37 | 0.5% | 30 | 0.6% | 7 | 0.3% |
| | White and Asian | 5 | 0.1% | 3 | 0.1% | 2 | 0.1% |
| | White and Black African | 3 | 0.0% | 2 | 0.0% | 1 | 0.0% |
| | White and Black Caribbean | 8 | 0.1% | 4 | 0.1% | 4 | 0.2% |
| | Missing | 281 | 3.9% | 196 | 4.1% | 85 | 3.7% |
| IMD Quintiles | 1 | 1,304 | 18.3% | 888 | 18.4% | 416 | 18.2% |
| | 2 | 1,321 | 18.5% | 941 | 19.4% | 380 | 16.6% |
| | 3 | 1,477 | 20.7% | 993 | 20.5% | 484 | 21.2% |
| | 4 | 1,514 | 21.3% | 999 | 20.6% | 515 | 22.5% |
| | 5 | 1,484 | 20.8% | 1,007 | 20.8% | 477 | 20.9% |
| | Missing | 23 | 0.3% | 11 | 0.2% | 12 | 0.5% |

| | | | | | | | | |
|---|---|---|---|---|---|---|---|---|
| Risk status | Clinical risk group (excl. severely immunosuppressed) | | 6,267 | 88.0% | 4,249 | 87.8% | 2,018 | 88.4% |
| | Severely immunosuppressed | | 394 | 5.5% | 270 | 5.6% | 124 | 5.4% |
| Carehome resident | | | 442 | 6.2% | 337 | 7.0% | 105 | 4.6% |
| Influenza vaccine 2023/2024 | | | 1,431 | 20.1% | 1,032 | 21.3% | 399 | 17.5% |
| COVID vaccine status | Spring 2023 booster* | None | 2,509 | 35.2% | 1,727 | 35.7% | 782 | 34.2% |
| | | 10-14 weeks | 290 | 4.1% | 205 | 4.2% | 85 | 3.7% |
| | | 15-19 weeks | 1,329 | 18.7% | 883 | 18.2% | 446 | 19.5% |
| | | 20-24 weeks | 1,891 | 26.5% | 1,211 | 25.0% | 680 | 29.8% |
| | | 25+ weeks | 1,104 | 15.5% | 813 | 16.8% | 291 | 12.7% |

*Sanofi/GSK booster or BA.4-5 bivalent, in addition to at least 2 prior doses waned for at least 3 months

Supplementary Table 5. Descriptive characteristics of cases and controls included in the analysis of the effectiveness of the bivalent BA.4-5 booster and monovalent XBB.1.5 booster given in autumn 2023 against hospitalisation amongst those aged 65 years and older, as presented in Table 2. Cases and controls by calendar week are shown in Supplementary Figure 1.

| | | Overall | | Controls | | Cases | |
|---|---|---|---|---|---|---|---|
| | | n | % | n | % | n | % |
| | Totals | 28,916 | 100% | 22,156 | 77% | 6,760 | 23% |
| Gender | Female | 14,515 | 50.2% | 11,292 | 51.0% | 3,223 | 47.7% |
| | Male | 13,853 | 47.9% | 10,324 | 46.6% | 3,529 | 52.2% |
| | Missing | 548 | 1.9% | 540 | 2.4% | 8 | 0.1% |
| | 65-69 | 3,137 | 10.8% | 2,518 | 11.4% | 619 | 9.2% |
| | 70-74 | 4,242 | 14.7% | 3,333 | 15.0% | 909 | 13.4% |
| | 75-79 | 5,727 | 19.8% | 4,396 | 19.8% | 1,331 | 19.7% |
| | 80-84 | 5,845 | 20.2% | 4,370 | 19.7% | 1,475 | 21.8% |
| | 85-89 | 5,376 | 18.6% | 3,994 | 18.0% | 1,382 | 20.4% |
| | 90+ | 4,589 | 15.9% | 3,545 | 16.0% | 1,044 | 15.4% |
| NHS Region | East of England | 2,426 | 8.4% | 1,851 | 8.4% | 575 | 8.5% |
| | London | 3,664 | 12.7% | 2,885 | 13.0% | 779 | 11.5% |
| | Midlands | 6,935 | 24.0% | 5,143 | 23.2% | 1,792 | 26.5% |
| | North East | 5,729 | 19.8% | 4,397 | 19.8% | 1,332 | 19.7% |
| | North West | 3,736 | 12.9% | 3,003 | 13.6% | 733 | 10.8% |
| | South East | 3,847 | 13.3% | 2,899 | 13.1% | 948 | 14.0% |
| | South West | 2,579 | 8.9% | 1,978 | 8.9% | 601 | 8.9% |
| Ethnicity | African | 97 | 0.3% | 75 | 0.3% | 22 | 0.3% |
| | Any other Asian background | 264 | 0.9% | 203 | 0.9% | 61 | 0.9% |
| | Any other Black background | 46 | 0.2% | 36 | 0.2% | 10 | 0.1% |
| | Any other White background | 1,293 | 4.5% | 991 | 4.5% | 302 | 4.5% |
| | Any other ethnic group | 259 | 0.9% | 205 | 0.9% | 54 | 0.8% |
| | Any other mixed background | 77 | 0.3% | 54 | 0.2% | 23 | 0.3% |
| | Bangladeshi or British Bangladeshi | 109 | 0.4% | 85 | 0.4% | 24 | 0.4% |
| | British, Mixed British | 24,114 | 83.4% | 18,419 | 83.1% | 5,695 | 84.2% |
| | Caribbean | 147 | 0.5% | 102 | 0.5% | 45 | 0.7% |
| | Chinese | 38 | 0.1% | 28 | 0.1% | 10 | 0.1% |
| | Indian or British Indian | 597 | 2.1% | 482 | 2.2% | 115 | 1.7% |
| | Irish | 383 | 1.3% | 301 | 1.4% | 82 | 1.2% |
| | Pakistani or British Pakistani | 309 | 1.1% | 254 | 1.1% | 55 | 0.8% |
| | White and Asian | 20 | 0.1% | 14 | 0.1% | 6 | 0.1% |
| | White and Black African | 17 | 0.1% | 14 | 0.1% | 3 | 0.0% |
| | White and Black Caribbean | 27 | 0.1% | 16 | 0.1% | 11 | 0.2% |
| | Missing | 1,119 | 3.9% | 877 | 4.0% | 242 | 3.6% |
| IMD Quintiles | 1 | 5,931 | 20.5% | 4,571 | 20.6% | 1,360 | 20.1% |
| | 2 | 5,519 | 19.1% | 4,260 | 19.2% | 1,259 | 18.6% |
| | 3 | 5,726 | 19.8% | 4,392 | 19.8% | 1,334 | 19.7% |
| | 4 | 5,918 | 20.5% | 4,506 | 20.3% | 1,412 | 20.9% |

| | | | | | | | | |
|---|---|---|---|---|---|---|---|---|
| | 5 | | 5,737 | 19.8% | 4,373 | 19.7% | 1,364 | 20.2% |
| | Missing | | 85 | 0.3% | 54 | 0.2% | 31 | 0.5% |
| Risk status | Clinical risk group (excl. severely immunosuppressed) | | 24,658 | 85.3% | 18,795 | 84.8% | 5,863 | 86.7% |
| | Severely immunosuppressed | | 1,900 | 6.6% | 1,449 | 6.5% | 451 | 6.7% |
| Carehome resident | | | 2,360 | 8.2% | 1,984 | 9.0% | 376 | 5.6% |
| Influenza vaccine 2023/2024 | | | 14,955 | 51.7% | 12,167 | 54.9% | 2,788 | 41.2% |
| COVID vaccine status* | Autumn 2023 booster | None | 14,578 | 50.4% | 10,227 | 46.2% | 4,351 | 64.4% |
| | BA.4-5 bivalent | 0-2 days | 139 | 0.5% | 121 | 0.5% | 18 | 0.3% |
| | | 3-8 days | 315 | 1.1% | 223 | 1.0% | 92 | 1.4% |
| | | 9-13 days | 270 | 0.9% | 206 | 0.9% | 64 | 0.9% |
| | | 2-4 weeks | 1,246 | 4.3% | 1,000 | 4.5% | 246 | 3.6% |
| | | 5-9 weeks | 2,221 | 7.7% | 1,914 | 8.6% | 307 | 4.5% |
| | | 10-14 weeks | 2,631 | 9.1% | 2,136 | 9.6% | 495 | 7.3% |
| | | 15+ weeks | 482 | 1.7% | 393 | 1.8% | 89 | 1.3% |
| | XBB.1.5 monovalent | 0-2 days | 150 | 0.5% | 124 | 0.6% | 26 | 0.4% |
| | | 3-8 days | 384 | 1.3% | 294 | 1.3% | 90 | 1.3% |
| | | 9-13 days | 362 | 1.3% | 290 | 1.3% | 72 | 1.1% |
| | | 2-4 weeks | 1,678 | 5.8% | 1,464 | 6.6% | 214 | 3.2% |
| | | 5-9 weeks | 2,685 | 9.3% | 2,292 | 10.3% | 393 | 5.8% |
| | | 10-14 weeks | 1,741 | 6.0% | 1,445 | 6.5% | 296 | 4.4% |
| | | 15+ weeks | 34 | 0.1% | 27 | 0.1% | 7 | 0.1% |

*In addition to at least two monovalent doses, waned for at least 3 months

Supplementary Table 6. Stratified and sensitivity analyses of the autumn 2023 booster against hospitalisation amongst those aged 65 years and older in England.

| Booster vaccine | Time since vaccination | Controls | Cases | OR | VE |
|---|---|---|---|---|---|
| Primary analysis (65 years and older) | | | | | |
| None | - | 10,227 | 4351 | Baseline | Baseline |
| Autumn 2023 booster* | 0-2 days | 245 | 44 | 0.34 (0.25-0.47) | 65.9 (52.5 to 75.5) |
| | 3-8 days | 517 | 182 | 0.69 (0.57-0.83) | 31.0 (17.1 to 42.5) |
| | 9-13 days | 496 | 136 | 0.59 (0.48-0.72) | 41.2 (28.0 to 52.1) |
| | 2-4 weeks | 2464 | 460 | 0.49 (0.44-0.56) | 50.6 (44.2 to 56.3) |
| | 5-9 weeks | 4206 | 700 | 0.54 (0.49-0.61) | 45.7 (39.4 to 51.3) |
| | 10-14 weeks | 3581 | 791 | 0.64 (0.57-0.71) | 36.5 (28.8 to 43.4) |
| | 15+ weeks | 420 | 96 | 0.86 (0.67-1.12) | 13.6 (-11.7 to 33.2) |
| Within those aged 75 years and older | | | | | |
| None | - | 7,084 | 3242 | Baseline | Baseline |
| Autumn 2023 booster* | 0-2 days | 190 | 39 | 0.37 (0.26-0.53) | 62.6 (46.7 to 73.8) |
| | 3-8 days | 407 | 145 | 0.66 (0.54-0.81) | 33.7 (18.5 to 46.1) |
| | 9-13 days | 382 | 110 | 0.59 (0.47-0.74) | 41.2 (26.2 to 53.2) |
| | 2-4 weeks | 1912 | 385 | 0.50 (0.44-0.58) | 49.6 (42.3 to 56.0) |
| | 5-9 weeks | 3229 | 572 | 0.54 (0.48-0.61) | 45.8 (38.7 to 52.0) |
| | 10-14 weeks | 2775 | 662 | 0.65 (0.57-0.73) | 35.5 (26.5 to 43.4) |
| | 15+ weeks | 326 | 77 | 0.87 (0.65-1.17) | 12.8 (-16.6 to 34.8) |
| Within those in a clinical risk group aged 18-64 years | | | | | |
| None | - | 3,550 | 904 | Baseline | Baseline |
| Autumn 2023 booster* | 0-2 days | 34 | 5 | 0.43 (0.16-1.13) | 57.1 (-13 to 83.7) |
| | 3-8 days | 78 | 15 | 0.66 (0.37-1.18) | 34.1 (-18.4 to 63.3) |
| | 9-13 days | 66 | 8 | 0.40 (0.19-0.86) | 59.8 (14.0 to 81.2) |
| | 2-4 weeks | 382 | 46 | 0.58 (0.41-0.83) | 41.5 (16.8 to 58.9) |
| | 5-9 weeks | 625 | 83 | 0.75 (0.56-1.00) | 24.9 (-0.4 to 43.9) |
| | 10-14 weeks | 410 | 68 | 0.93 (0.67-1.28) | 7.4 (-27.6 to 32.8) |
| | 15+ weeks | 38 | 7 | n too small | n too small |
| With adjustment for prior infection (65 years and older) | | | | | |
| None | - | 10,227 | 4351 | Baseline | Baseline |
| Autumn 2023 booster* | 0-2 days | 245 | 44 | 0.37 (0.26-0.51) | 63.2 (48.7 to 73.6) |
| | 3-8 days | 517 | 182 | 0.70 (0.58-0.84) | 30.4 (16.3 to 42.1) |
| | 9-13 days | 496 | 136 | 0.59 (0.48-0.72) | 41.5 (28.2 to 52.3) |
| | 2-4 weeks | 2464 | 460 | 0.50 (0.44-0.57) | 50.0 (43.5 to 55.7) |
| | 5-9 weeks | 4206 | 700 | 0.55 (0.49-0.61) | 45.4 (39.1 to 51.0) |
| | 10-14 weeks | 3581 | 791 | 0.64 (0.57-0.72) | 36.2 (28.4 to 43.2) |
| | 15+ weeks | 420 | 96 | 0.87 (0.68-1.13) | 12.6 (-13.2 to 32.5) |
| Without adjustment for seasonal (2023/2024) influenza vaccination (65 years and older) | | | | | |
| None | - | 10,227 | 4351 | Baseline | Baseline |
| Autumn 2023 booster* | 0-2 days | 245 | 44 | 0.34 (0.25-0.47) | 65.8 (52.5 to 75.4) |
| | 3-8 days | 517 | 182 | 0.69 (0.58-0.83) | 30.8 (17.3 to 42.2) |
| | 9-13 days | 496 | 136 | 0.59 (0.48-0.72) | 41.1 (28.2 to 51.8) |

| | | | | | |
|---|---|---|---|---|---|
| | 2-4 weeks | 2464 | 460 | 0.49 (0.44-0.55) | 50.5 (44.5 to 55.9) |
| | 5-9 weeks | 4206 | 700 | 0.54 (0.49-0.60) | 45.6 (39.7 to 50.8) |
| | 10-14 weeks | 3581 | 791 | 0.64 (0.57-0.71) | 36.4 (29.1 to 42.9) |
| | 15+ weeks | 420 | 96 | 0.87 (0.67-1.12) | 13.5 (-11.6 to 32.9) |
| Including all hospital admissions with at least a 2 day length of stay (65 years and older) | | | | | |
| None | - | 44,116 | 7955 | Baseline | Baseline |
| Autumn 2023 booster* | 0-2 days | 835 | 80 | 0.46 (0.36-0.58) | 53.9 (41.8 to 63.6) |
| | 3-8 days | 2083 | 296 | 0.67 (0.59-0.76) | 32.9 (23.5 to 41.0) |
| | 9-13 days | 1914 | 228 | 0.59 (0.51-0.68) | 41.2 (32.0 to 49.1) |
| | 2-4 weeks | 8906 | 786 | 0.51 (0.46-0.55) | 49.4 (44.9 to 53.6) |
| | 5-9 weeks | 15824 | 1454 | 0.57 (0.53-0.61) | 43.5 (39.3 to 47.4) |
| | 10-14 weeks | 15262 | 2067 | 0.71 (0.66-0.76) | 28.9 (23.8 to 33.7) |
| | 15+ weeks | 2996 | 389 | 0.85 (0.74-0.96) | 15.5 (4.0 to 25.6) |
| Comparing to not boosted, regardless of past vaccination | | | | | |
| None | - | 11,015 | 4669 | Baseline | Baseline |
| Autumn 2023 booster* | 0-2 days | 245 | 45 | 0.35 (0.25-0.48) | 65.3 (51.9 to 75.0) |
| | 3-8 days | 522 | 184 | 0.69 (0.57-0.82) | 31.4 (17.7 to 42.9) |
| | 9-13 days | 500 | 137 | 0.58 (0.48-0.71) | 41.7 (28.5 to 52.4) |
| | 2-4 weeks | 2480 | 461 | 0.49 (0.43-0.55) | 51.4 (45.1 to 56.9) |
| | 5-9 weeks | 4230 | 704 | 0.53 (0.48-0.60) | 46.5 (40.4 to 52.0) |
| | 10-14 weeks | 3598 | 796 | 0.63 (0.56-0.70) | 37.5 (29.9 to 44.2) |
| | 15+ weeks | 424 | 97 | 0.85 (0.66-1.10) | 14.6 (-10.4 to 33.9) |

*Of the bivalent BA.4-5 booster or the monovalent XBB.1.5 booster

Supplementary Table 7. Descriptive characteristics of cases and controls included in the analysis of the effectiveness of the bivalent BA.4-5 booster and monovalent XBB.1.5 booster given in autumn 2023 against hospitalisation amongst those aged 18-64 in a clinical risk group, as presented in Supplementary Table 6.

| | | Overall | | Controls | | Cases | |
|---|---|---|---|---|---|---|---|
| | | n | % | n | % | n | % |
| | Totals | 6,319 | 100% | 5,183 | 82% | 1,136 | 18% |
| Gender | Female | 3,292 | 52.1% | 2,662 | 51.4% | 630 | 55.5% |
| | Male | 2,865 | 45.3% | 2,361 | 45.6% | 504 | 44.4% |
| | Missing | 162 | 2.6% | 160 | 3.1% | 2 | 0.2% |
| | 18-19 | 41 | 0.6% | 33 | 0.6% | 8 | 0.7% |
| | 20-24 | 145 | 2.3% | 125 | 2.4% | 20 | 1.8% |
| | 25-29 | 178 | 2.8% | 144 | 2.8% | 34 | 3.0% |
| | 30-34 | 251 | 4.0% | 209 | 4.0% | 42 | 3.7% |
| | 35-39 | 309 | 4.9% | 265 | 5.1% | 44 | 3.9% |
| | 40-44 | 424 | 6.7% | 343 | 6.6% | 81 | 7.1% |
| | 45-49 | 554 | 8.8% | 469 | 9.0% | 85 | 7.5% |
| | 50-54 | 953 | 15.1% | 779 | 15.0% | 174 | 15.3% |
| | 55-59 | 1,437 | 22.7% | 1,181 | 22.8% | 256 | 22.5% |
| | 60-64 | 2,027 | 32.1% | 1,635 | 31.5% | 392 | 34.5% |
| NHS Region | East of England | 449 | 7.1% | 369 | 7.1% | 80 | 7.0% |
| | London | 873 | 13.8% | 715 | 13.8% | 158 | 13.9% |
| | Midlands | 1,510 | 23.9% | 1,219 | 23.5% | 291 | 25.6% |
| | North East | 1,214 | 19.2% | 1,020 | 19.7% | 194 | 17.1% |
| | North West | 989 | 15.7% | 834 | 16.1% | 155 | 13.6% |
| | South East | 766 | 12.1% | 620 | 12.0% | 146 | 12.9% |
| | South West | 518 | 8.2% | 406 | 7.8% | 112 | 9.9% |
| Ethnicity | African | 112 | 1.8% | 84 | 1.6% | 28 | 2.5% |
| | Any other Asian background | 108 | 1.7% | 88 | 1.7% | 20 | 1.8% |
| | Any other Black background | 53 | 0.8% | 46 | 0.9% | 7 | 0.6% |
| | Any other White background | 300 | 4.7% | 248 | 4.8% | 52 | 4.6% |
| | Any other ethnic group | 101 | 1.6% | 86 | 1.7% | 15 | 1.3% |
| | Any other mixed background | 38 | 0.6% | 30 | 0.6% | 8 | 0.7% |
| | Bangladeshi or British Bangladeshi | 64 | 1.0% | 60 | 1.2% | 4 | 0.4% |
| | British, Mixed British | 4,841 | 76.6% | 3,965 | 76.5% | 876 | 77.1% |
| | Caribbean | 66 | 1.0% | 50 | 1.0% | 16 | 1.4% |
| | Chinese | 22 | 0.3% | 17 | 0.3% | 5 | 0.4% |
| | Indian or British Indian | 173 | 2.7% | 144 | 2.8% | 29 | 2.6% |
| | Irish | 47 | 0.7% | 38 | 0.7% | 9 | 0.8% |
| | Pakistani or British Pakistani | 178 | 2.8% | 151 | 2.9% | 27 | 2.4% |
| | White and Asian | 20 | 0.3% | 17 | 0.3% | 3 | 0.3% |
| | White and Black African | 10 | 0.2% | 8 | 0.2% | 2 | 0.2% |
| | White and Black Caribbean | 19 | 0.3% | 14 | 0.3% | 5 | 0.4% |
| | Missing | 167 | 2.6% | 137 | 2.6% | 30 | 2.6% |

| | | | | | | | |
|---|---|---|---|---|---|---|---|
| IMD Quintiles | 1 | 1,968 | 31.1% | 1,663 | 32.1% | 305 | 26.8% |
| | 2 | 1,402 | 22.2% | 1,149 | 22.2% | 253 | 22.3% |
| | 3 | 1,128 | 17.9% | 916 | 17.7% | 212 | 18.7% |
| | 4 | 952 | 15.1% | 762 | 14.7% | 190 | 16.7% |
| | 5 | 825 | 13.1% | 661 | 12.8% | 164 | 14.4% |
| | Missing | 44 | 0.7% | 32 | 0.6% | 12 | 1.1% |
| Risk status | Clinical risk group (excl. severely immunosuppressed) | 5,647 | 89.4% | 4,657 | 89.9% | 990 | 87.1% |
| | Severely immunosuppressed | 672 | 10.6% | 526 | 10.1% | 146 | 12.9% |
| Influenza vaccine 2023/2024 | | 2,314 | 36.6% | 2,005 | 38.7% | 309 | 27.2% |
| Autumn 2023 booster* | None | 4,454 | 70.5% | 3,550 | 68.5% | 904 | 79.6% |
| | 0-2 days | 39 | 0.6% | 34 | 0.7% | 5 | 0.4% |
| | 3-8 days | 93 | 1.5% | 78 | 1.5% | 15 | 1.3% |
| | 9-13 days | 74 | 1.2% | 66 | 1.3% | 8 | 0.7% |
| | 2-4 weeks | 428 | 6.8% | 382 | 7.4% | 46 | 4.0% |
| | 5-9 weeks | 708 | 11.2% | 625 | 12.1% | 83 | 7.3% |
| | 10-14 weeks | 478 | 7.6% | 410 | 7.9% | 68 | 6.0% |
| | 15+ weeks | 45 | 0.7% | 38 | 0.7% | 7 | 0.6% |

*In addition to at least two monovalent doses, waned for at least 3 months

Supplementary Figure 1. Cases and controls by calendar week in the study period for (a) adults aged 18 to 49 years (1 and Supplementary Table 1); (b) adults aged 50 to 63 years (Table 1 and Supplementary Table 2); (c) adults aged 65 years and older (Table 1 and Supplementary Table 3); (d) adults aged 75 years and older (Table 1 and Supplementary Table 4); (e) adults aged 65 years and older included in the analysis of the autumn 2023 booster (Table 2 and Supplementary Table 5).

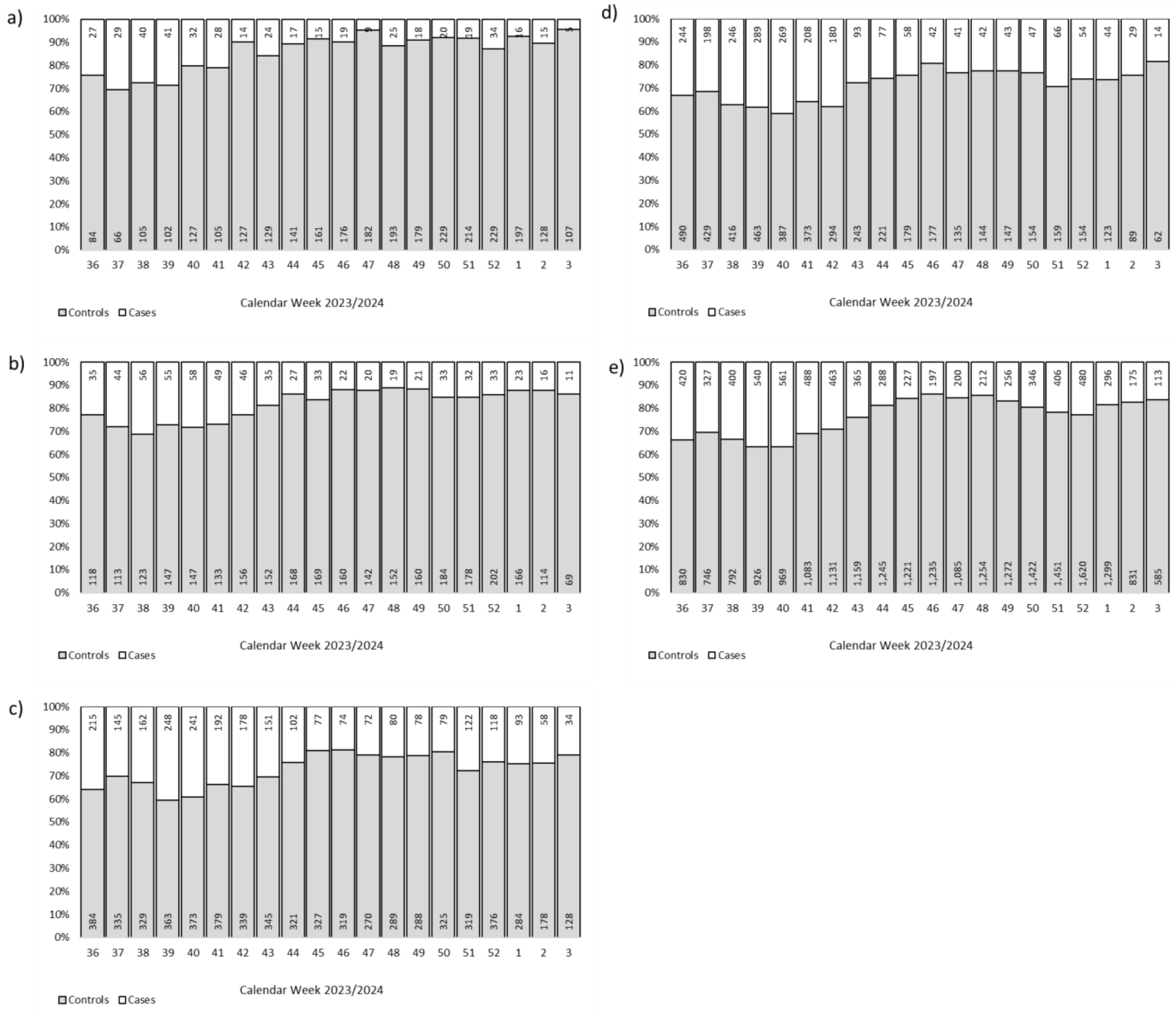

Supplementary Figure 2. Number of cases of XBB, XBB.1.5, XBB.1.6, EG.5.1 and JN.1 by calendar week over time included in the analysis of vaccine effectiveness by variant as presented in Table 3.

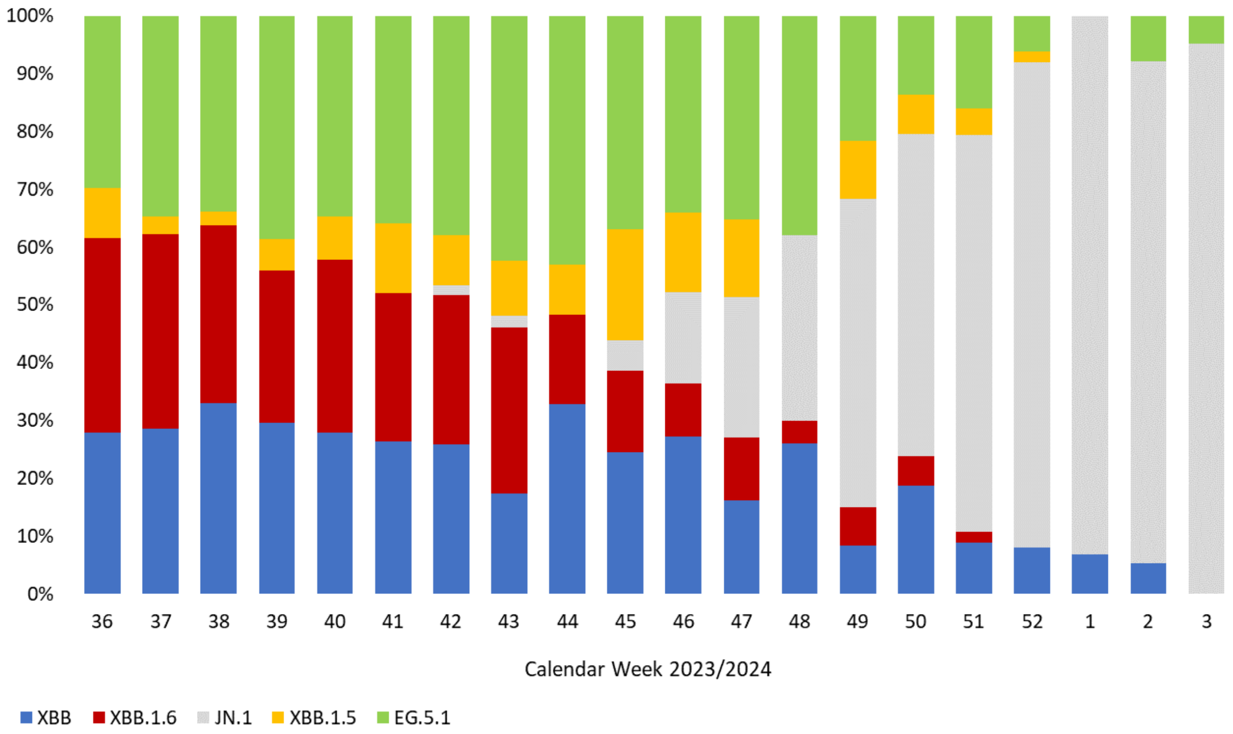

Supplementary Figure 3. The distribution of time since booster vaccination by variant of cases included in the analysis of vaccine effectiveness by variant presented in Table 3.

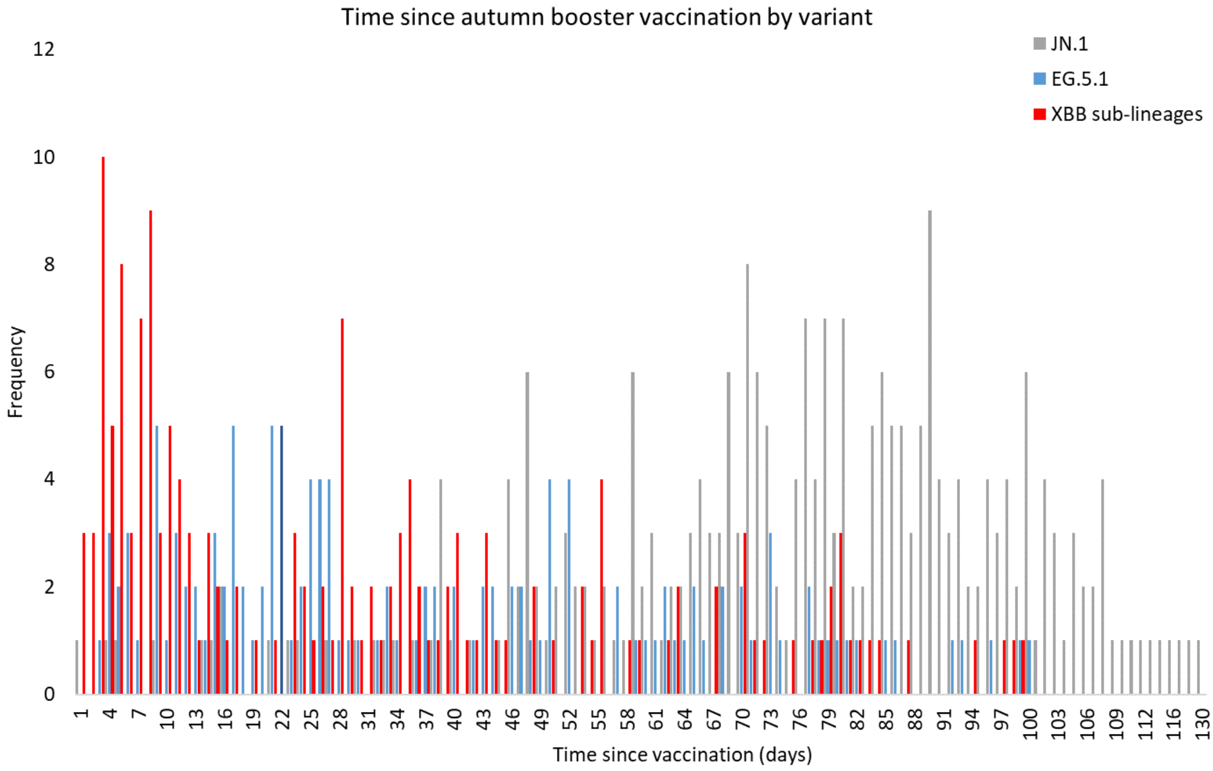
